# Suspected COVID-19 in primary care: how GP records contribute to understanding differences in prevalence by ethnicity

**DOI:** 10.1101/2020.05.23.20101741

**Authors:** Sally A Hull, Crystal Williams, Mark Ashworth, Chris Carvalho, Kambiz Boomla

**Affiliations:** Institute of Population Health Sciences, Queen Mary University of London, 58 Turner Street, London E1 2AB; School of Population Health and Environmental Sciences, King’s College London, Guy’s Campus, London SE1 1UL.

**Keywords:** Primary care, ethnicity, multi-morbidity, COVID-19

## Abstract

**Background:** The first wave of the London COVID-19 epidemic peaked in April 2020. Attention initially focussed on severe presentations, intensive care capacity, and the timely supply of equipment. General practice has seen a rapid take up of technology to allow virtual consultations, enabling the management of mild and moderate community cases.

**Aim:** To quantify the prevalence and time-course of suspected COVID-19 presenting to general practices during the London epidemic. To report disease prevalence by ethnic group, and explore how far differences by ethnicity can be explained by data in the electronic health record (EHR).

**Design and Setting:** Cross-sectional study using anonymised data from the primary care records of 1.3 million people registered with 157 practices in four adjacent east London clinical commissioning groups (CCGs). The study area includes 48% of people from ethnic minorities and is in the top decile of social deprivation in England.

**Method:** Suspected COVID-19 cases were identified using SNOMED codes.

Explanatory variables included age, gender, self-reported ethnicity and measures of social deprivation. Clinical factors included 16 long-term conditions, latest body mass index and smoking status.

**Results:** There were 8,985 suspected COVID-19 cases. Ethnicity recording was 78% complete. Univariate analysis showed a two-fold increase in odds of infection for South Asian and Black adults compared to White. In a fully adjusted analysis, including clinical factors, the odds were: South Asian OR 1.93 (95% CI = 1.83 to 2.04) Black OR 1.47 (95% CI 1.38 to 1.57)

**Conclusions:** Using data in GP records Black and south Asian ethnicity remain as predictors of community cases of COVID-19, with levels of risk similar to hospital admission cases. Further understanding of these differences requires social and occupational data.

**What is known:** South Asian and Black ethnicity patients are at increased risk of hospital admission, intensive care admission and death from COVID-19 infection, compared with White ethnicity patients. However, little is known about the pattern of COVID-19 infection in a primary care population.

**What this study adds:** South Asian and Black ethnicity patients are at increased risk of a suspected COVID-19 diagnosis in primary care, even after adjustment for other factors which might also be associated with increased risk.

Multimorbidity, increasing obesity and social deprivation are also strongly associated with increased risk of a suspected COVID-19 diagnosis.

Primary care recording of suspected COVID-19 diagnoses closely mirrors COVID-19 test positivity reported by the national testing scheme. Recording rates of suspected COVID-19 may provide an early warning system for any future upward trends in transmission rates.

## Introduction

The rapid worldwide spread of COVID-19 in early 2020, from its origin in Wuhan China, (1) led the World Health Organisation to declare a pandemic on 11 March 2020.(2)

In the UK early attention focussed on hospital presentations and intensive care capacity, the timely supply of equipment, and latterly the increasing death rate in care home settings. (3-5) Community testing – which forms part of standard public health test and quarantine policy - ceased in England on 12 March, (6) hence the extent of asymptomatic and milder symptomatic cases within community settings remains unknown. Early evidence from testing among passengers on cruise ships, (7) suggests that 18% of infected people have no symptoms. The figures are likely to be higher in populations with a younger demographic profile.

Up to mid-April 2020 London has had the highest age standardised mortality rate for deaths in the UK reported as Corona virus, with 85.7 deaths per 100,000 population (compared to 36.6/100,000 in England). Three of the four east London localities in this study had death rates in the top 5 for London boroughs. (Newham 144.3, Hackney 127.4, Tower Hamlets 122.9) (8) Data from ONS, and the OpenSAFELY study indicate that mortality rates in the most deprived areas of England were almost twice as high as those in the least deprived areas, and that males had higher death rates than females. (8, 9)

From an early stage in the UK epidemic people with COVID-19 like symptoms were advised not to attend their general practice in person, and to use online or phone contact with NHS 111. (10) Throughout general practice there was rapid take up of technological solutions to facilitate a shift to telephone and video consultations, which enabled GPs to manage community cases – despite the national failure to share COVID-19 test results offered by drive-through or home-based tests. (11, 12) Practices worked collectively to provide separate locations for the necessary physical examinations of people with suspected COVID-19 cases and for those with other medical problems. (13)

Concern has been raised about the higher case fatality rate of Black, Asian, and minority ethnic (BAME) patients in intensive care units (14) and the disproportionate numbers of deaths of health and social care workers from these groups.

The population of east London includes 48% of people from minority ethnic backgrounds. Hence this geographical area is well placed to examine whether Black and south Asian populations are over-represented in the population consulting their GP practice with suspected COVID-19 symptoms, and to explore health related causes of these differences.

*Study aim:*

a. To identify the numbers of clinically suspected COVID-19 cases recorded by practices through the peak of the London epidemic in February to end of April 2020.
b. To examine whether there is an excess of clinically suspected cases among the major ethnic minority groups, and how far this can be accounted for by differences in demographic status, or by differences in the burden of long-term conditions.

## Methods

### Design and setting

A cross sectional study using primary care electronic health data from 1.2 million adult patients registered at 157 general practices in the four geographically contiguous east London Clinical Commissioning Groups of Newham, Tower Hamlets, City & Hackney and Waltham Forest. In the 2011 UK Census 48% of the population in these CCGs was recorded as being of non-White ethnic origin, (15) and the English indices of deprivation 2015 show that all four feature in the top decile of the most socially deprived boroughs in England. (16)

### Data sources

The study population included all adults (>18 years) registered at the 157 practices at the start of the study period, 1^st^ January to end April 2020. Data were extracted on secure N3 terminals from EMIS Web, used by (157/162) practices in the study area. All data was extracted and managed according to UK NHS information governance requirements.

### Data extracted

Sociodemographic variables included age, gender and self-reported ethnicity captured at the time of registration or during routine consultations. Ethnic categories were based on the 18 categories of the UK 2011 census and were combined into four groups reflecting the study population: White (British, Irish, other White), Black (Black African, Black Caribbean, Black British, other Black and mixed Black), South Asian (Bangladeshi, Pakistani, Indian, Sri Lankan, British Asian, other Asian or mixed Asian), and Other (Chinese, Arab, any other ethnic group). Individuals of mixed ethnicity were grouped with their parent ethnic minority for the purposes of this study. (17) Patients with other, unknown, or missing ethnicity were not reported in the final analysis. The English indices of deprivation (IMD) 2015 score was used as a measure of social deprivation. (16) The IMD score for each patient was mapped to the patient lower super output area (LSOA) of residence to derive internal and national quintiles for the study population.

Clinical measures included the COVID-19 SNOMED codes, which were supplied to GP computer systems from 6 February. (18) Codes for cough, fever, upper respiratory tract infection, flu like illness and lower respiratory tract infection were also extracted. These may have been used for symptomatic cases prior to the release of the COVID-19 codes, and potentially during the course of the epidemic.

To assess the burden of long term conditions (LTCs) in the study population we extracted diagnostic data on 16 LTCs which form part of the UK Quality and Outcomes Framework (QOF), using the earliest recorded diagnostic code prior to the start of the study, based on version 44 of the QOF business rule set. (19) The conditions included were: asthma, chronic obstructive pulmonary disease (COPD), atrial fibrillation, heart failure, hypertension, coronary heart disease (CHD), peripheral arterial disease (PAD), stroke and transient ischaemic attack, chronic kidney disease (CKD), diabetes, dementia, depression, epilepsy, learning disabilities, serious mental illness, and cancer. The total count of these QOF LTCs per person was used as the principal measure of multi-morbidity in the adult population. (20, 21) The effect of different individual LTCs was explored in a sensitivity analysis.

We extracted routine clinical data on body mass index (BMI) and smoking status as the latest recorded codes prior to the start of the study period. BMI values were categorised as underweight, normal, overweight, obese and morbidly obese.

Data on test-confirmed COVID-19 cases across London and the study CCGs were obtained from the UK Government Digital Service website. (22)

### Statistical Analysis

Our primary outcome measure was prevalence of suspected COVID-19 recorded in the EHR. All statistical analysis was undertaken in Stata version 16.1 (College Station, TX: StataCorp LP.) We fitted logistic, mixed effect models, nesting patients within practices. Both univariate and multivariate models were fitted. The effect of ethnicity on the likelihood of suspected COVID-19 presentation was examined, adjusting for differences in demographic and clinical factors including long term conditions and BMI.

Sensitivity analyses were undertaken using individual co-morbidities in place of counts of conditions.

### Information governance and ethics

The clinical effectiveness group is the data processor, and the General Practices in the four CCGs are the data controllers. CEG has the written consent of all practices in the study area to use pseudonymised patient data for audit and research for patient benefit. The researchers adhere to the data protection principles of the Data Protection Act 2018, and all data was managed according to UK NHS information governance requirements. All outputs were in the form of aggregate patient data.

The NHS Health Research Authority toolkit (http://www.hra-decisiontools.org.uk/ethics/) identified that Research Ethics Approval was not required for this project as all data is pseudonomised and presented in aggregate form. This was confirmed by the Chair of the North East London Strategic Information Governance Network.

### Patient and public Involvement

Patients and members of the public were not involved in the design or reporting of this study.

## Results

Primary care data from the records of 1,257,130 adult patients registered at 157 practices was available for analysis. Among this population 8,985 (0.7%) had a record of suspected COVID-19 between 14 February and 31 April 2020, and 35,022 (2.8%) had a code for URTI or LRTI infection between 1^st^ January and 31 April.

Figure 1. compares the daily count of test-positive COVID-19 cases across all of London, with those in the four study CCGs, demonstrating that the distribution of test-positive cases follows a similar time course. The daily count of test-positive COVID-19 cases in the study area is compared with GP codes for suspected COVID-19 cases in Figure 2, demonstrating a similar distribution, but threefold greater numbers. Figure 3. shows the daily counts of URTI/LRTI cases from 1 January to 30 April compared with suspected COVID-19 cases. This demonstrates the seasonal decline of URTI/LRTI cases in April, possibly magnified by social distancing.

**Figure 1.**
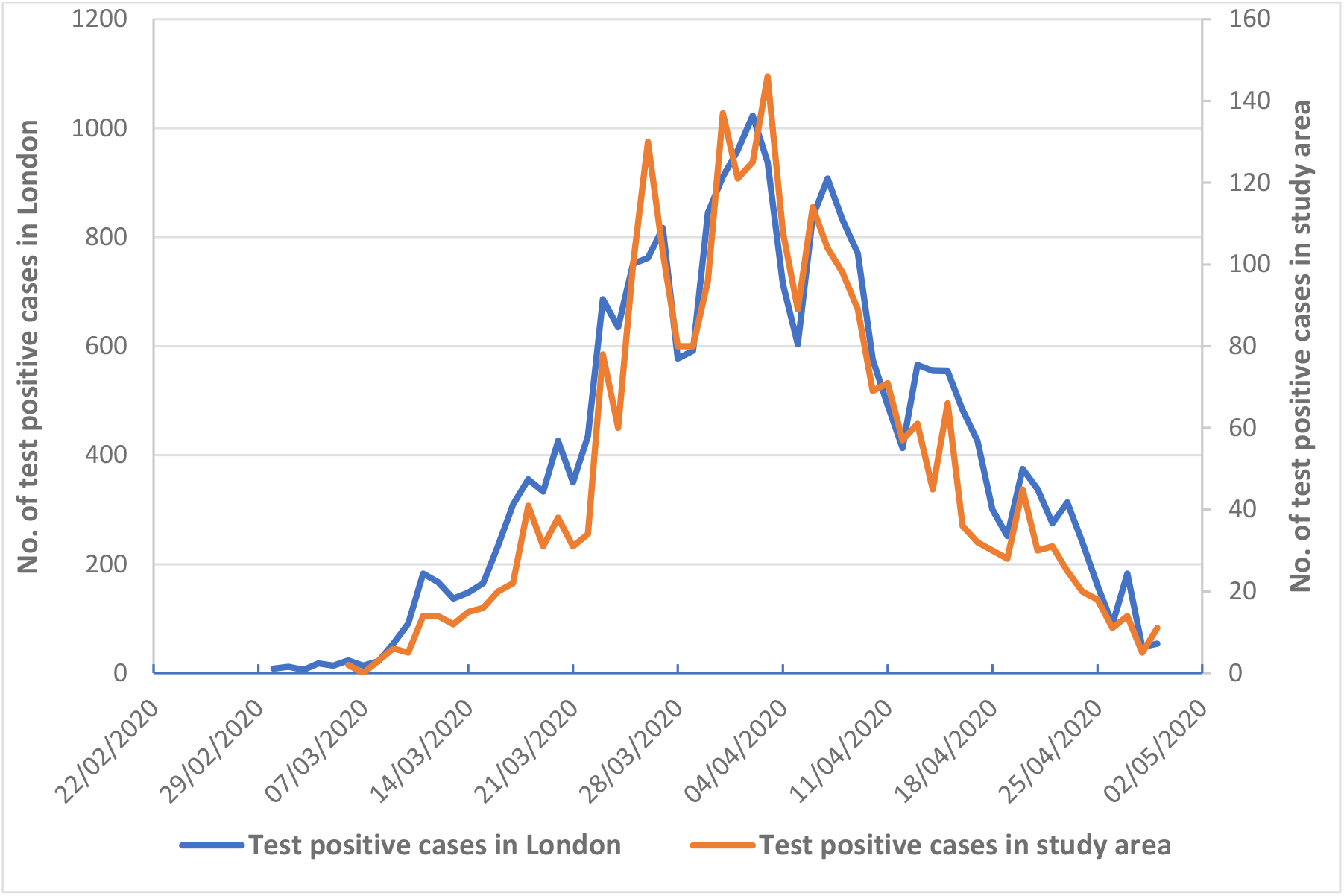
Comparison of test-positive cases in the whole of London, and those in the east London study area: February to April 2020.

The characteristics of patients with and without COVID-19 symptoms are shown in Table 1. Ethnicity recording was 78% complete. Univariate analysis demonstrates that compared to White adults south Asian and Black adults had almost twice the odds of infection (South Asian 1.98 (95% CI 1.86 to 2.09) Black 1.88 (95% CI 1.77 to 2.0)). 11.3% of adults had more than one long term condition, and smoking prevalence was 17.3%, similar to the England average of 16.9%. Valid BMI results were available for 80% of cases.

The univariate analysis (Table 1) shows a two-fold increase in odds by social deprivation, with 88% of the population falling into the 4^th^ and 5^th^ (most deprived) national quintiles of the English IMD scores. There is a steep increase of odds associated with increasing numbers of LTCs and with BMI categories. All LTCs were associated with increased odds, the odds ratio for dementia (OR 7.37) may reflect the population living in residential and nursing homes.

**Table 1.** Characteristics of those with and without suspected COVID-19 codes to the end of April 2020. (Includes 1,257,136 patients aged ≥ 18 years from 165 practices)

|  | Suspected<br>COVID-19 (%) | Without suspected<br>COVID-19 (%) | Univariate OR<br>(95%CI) |
| --- | --- | --- | --- |
| **Total** | 8,985 | 1,248,152 |  |
| **CCG** |  |  |  |
| Tower Hamlets | 2,558 (28.5) | 292,653 (23.5) |  |
| Newham | 2,732 (30.4) | 377,171 (30.2) |  |
| City & Hackney | 2,674 (29.8) | 351,060 (28.1) |  |
| Waltham Forest | 1,021 (11.4) | 227,268 (18.2) |  |
| **Age** |  |  |  |
| 18-49 (ref) | 5,134 (57.1) | 926,886 (74.3) |  |
| 50-69 | 2,723 (30.3) | 235,616 (18.9) | 2.18 (2.08-2.29) |
| over 70 | 1,128 (12.6) | 85,650 (6.9) | 2.45(2.29-2.62) |
| **Sex** |  |  |  |
| Male (ref) | 3,982 (44.3) | 632,082 (50.6) |  |
| Female | 5,003 (55.7) | 616,070 (49.4) | 1.28 (1.22-1.33) |
| **Ethnicity** |  |  |  |
| White (ref) | 2,890 (32.2) | 476,302 (38.2) |  |
| South Asian | 2,859 (31.8) | 259,464 (20.8) | 1.98 (1.86-2.09) |
| Black | 1,642 (18.3) | 153,240 (12.3) | 1.88 (1.77-2.00) |
| Other | 594 (6.6) | 78,454 (6.3) | 1.24 (1.13-1.35) |
| Not Stated/Missing | 1,000 (11.1) | 280,692 (22.5) | 0.64 (0.60-0.69) |
| **National IMD 2015 quintiles** |  |  |  |
| 1 least deprived (ref) | 30 (0.3) | 8964 (0.7) |  |
| 2 | 96 (1.1) | 24029 (1.9) | 1.35 (0.88-2.06) |
| 3 | 485 (5.4) | 99395 (8.0) | 1.22 (0.83-1.79) |
| 4 | 3557 (39.6) | 541773 (43.4) | 1.53 (1.05-2.23) |
| 5 most deprived | 4807 (53.5) | 560245 (44.9) | 1.88 (1.29-2.74) |
| Missing | 10 (0.1) | 13746 (1.1) | 0.21 (0.10- 0.43) |
| **BMI** |  |  |  |
| Normal weight (18.5 to <25) (ref) | 2,528 (28.1) | 431,279 (34.6) |  |
| Underweight (<18.5) | 200 (2.2) | 39,067 (3.1) | 0.85 (0.73-1.00) |
| Overweight (25 to <30) | 2,770 (30.8) | 299,136 (24.0) | 1.60 (1.52 - 1.69) |
| Obese (30 to <40) | 2,451 (27.3) | 169,982 (13.6) | 2.49 (2.35-2.63) |
| Morbidly obese ($\geq 40$) | 483 (5.4) | 23,717 (1.9) | 3.48 (3.15-3.84) |
| Out of range/Unknown | 553 (6.2) | 284,971 (22.8) | 0.33 (0.30-0.36) |
| **QOF long term conditions** |  |  |  |
| 0 (ref) | 3,740 (41.6) | 881,460 (70.6) |  |
| 1 | 2,461 (27.4) | 226,961 (18.2) | 2.41 (2.29-2.54) |
| 2 | 1,350 (15.0) | 81,093 (6.6) | 3.75 (3.52-3.99) |
| 3 | 690 (7.7) | 33,497 (2.7) | 4.6 (4.25-5.02) |
| 4+ | 744 (8.3) | 25,141 (2.0) | 6.5 (6.00-7.05) |
| **Current smoker** | 1,047 (11.7) | 217,396 (17.4) | 0.60 (0.56-0.63) |
| **Asthma** | 1,512 (16.8) | 111,641 (8.9) | 1.92 (1.81-2.03) |
| **Atrial Fibrillation** | 248 (2.8) | 10,299 (0.8) | 3.16 (2.78-3.59) |
| **Cancer** | 429 (4.8) | 22,989 (1.8) | 2.50 (2.26-2.75) |
| **Coronary heart disease** | 504 (5.6) | 23,114 (1.9) | 2.98 (2.72 -3.26) |
| **Chronic kidney disease (3-5)** | 716 (8.0) | 32,203 (2.6) | 3.11 (2.88-3.37) |
| **COPD** | 331 (3.7) | 14,467 (1.2) | 2.92 (2.61-3.26) |
| **Dementia** | 258 (2.9) | 4,442 (0.36) | 7.37 (6.48-8.39) |
| **Depression** | 1,811 (20.2) | 121,290 (9.7) | 2.15 (2.04-2.27) |
| **Diabetes** | 1,696 (18.9) | 79,445 (6.4) | 3.31 (3.13-3.49) |
| **Epilepsy** | 157 (1.8) | 10,321 (0.8) | 2.00 (1.70 - 2.34) |
| **Heart Failure** | 234 (0.7) | 8,039 (0.6) | 3.75 (3.28-4.28) |
| **Hypertension** | 2,290 (25.5) | 131,318 (10.5) | 2.85 (2.71-2.99) |
| **Learning disability** | 70 (0.8) | 4,660 (0.4) | 1.89 (1.49-2.40) |
| **Severe Mental Illness** | 250 (2.8) | 17,322 (1.4) | 1.88 (1.65-2.13) |
| **Peripheral arterial disease** | 87 (1.0) | 3,608 (0.3) | 3.00 (2.41-3.71) |
| **Stroke TIA** | 284 (3.2) | 11,514 (0.9) | 3.24 (2.87-3.65) |

CCG = clinical commissioning group, IMD = Index of Multiple Deprivation, OR = odds ratio, BMI = body mass index, QOF = Quality and Outcomes Framework, ref = reference.

Table 2 shows the multivariate model for predictors of COVID-19 for adults aged ≥ 18 years. This is divided into two sections, the first showing adjustment for age, gender and social deprivation, and the second showing a fully adjusted model including the clinical predictors. For these models internal quintiles of deprivation were used rather than national quintiles. The fully adjusted model shows that compared to White adults, south Asian adults still had nearly twice the odds of infection OR 1.93 (95% CI 1.83 to 2.04), the OR for Black adults had diminished to 1. 47 (95% CI 1.38 to 1.57). There is a steep gradient of odds associated with increasing numbers of LTCs and categories of BMI, however these factors do not have much explanatory effect on the prevalence of disease by ethnicity. The effect of social deprivation on the odds of infection was reduced in the fully adjusted model, OR 1.26 (95%CI 1.17 to 1.37).

**Table 2.** Multivariate model for predictors of COVID-19 for adults aged > 18 years (n=1,257,137 cases contributing to the model)

|  |  | Model 1.<br>Demographic factors |  |  | Model 2.<br>Demographic and Clinical factors |  |  |
| --- | --- | --- | --- | --- | --- | --- | --- |
|  |  | OR<sup>a</sup> | 95% CI | P Value | OR<sup>a</sup> | 95% CI | P Value |
| **Sex** | Male (ref) | 1.00 |  |  | 1.00 |  |  |
|  | female | 1.25 | (1.20 to 1.31) | <0.001 | 1.17 | (1.12 to 1.22) | <0.001 |
| **Age bands (years)** | 18-49 (ref) | 1.00 |  |  | 1.00 |  |  |
|  | 50-69 | 2.14 | (2.03 to 2.24) | <0.001 | 1.30 | (1.23 to 1.37) | <0.001 |
| | $\geq 69$ | 2.57 | (2.40 to 2.74) | <0.001 | 1.25 | (1.16 to 1.35) | <0.001 |
| **Ethnicity<sup>c</sup>** | White (ref) | 1.00 |  |  | 1.00 |  |  |
|  | South Asian | 2.06 | (1.94 to 2.18) | <0.001 | 1.93 | (1.83 to 2.04) | <0.001 |
|  | Black | 1.66 | (1.56 to 1.77) | <0.001 | 1.47 | (1.38 to 1.57) | <0.001 |
| **Internal IMD 2015 quintiles<sup>c</sup>** | 1 least deprived (ref) | 1.00 |  |  | 1.00 |  |  |
|  | 2 | 1.24 | (1.14 to 1.33) | <0.001 | 1.18 | (1.09 to 1.28) | <0.001 |
|  | 3 | 1.23 | (1.13 to 1.32) | <0.001 | 1.16 | (1.07 to 1.25) | <0.001 |
|  | 4 | 1.32 | (1.22 to 1.43) | <0.001 | 1.21 | (1.17 to 1.37) | <0.001 |
|  | 5 most deprived | 1.40 | (1.29 to 1.51) | <0.001 | 1.26 | (1.17 to 1.37) | <0.001 |
| **QOF long term conditions** | 0 (ref) |  |  |  | 1.00 |  |  |
|  | 1 |  |  |  | 1.77 | (1.67 to 1.87) | <0.001 |
|  | 2 |  |  |  | 2.28 | (2.13 to 2.45) | <0.001 |
|  | 3 |  |  |  | 2.60 | (2.37 to 2.85) | <0.001 |
| | $\geq 4$ | | | | 3.67 | (3.33 to 4.03) | <0.001 |
| **BMI<sup>d</sup>, Kg/m<sup>2</sup>** | Normal weight (18.5 to <25) (ref) |  |  |  | 1.00 |  |  |
|  | Underweight (<18.5) |  |  |  | 0.84 | (0.73 to 0.97) | 0.02 |
|  | Overweight (25 to <30) |  |  |  | 1.31 | (1.24 to 1.38) | <0.001 |
|  | Obese (30 to <35) |  |  |  | 1.73 | (1.63 to 1.84) | <0.001 |
| | Morbidly Obese ($\geq 40$) | | | | 2.20 | (2.01 to 2.47) | <0.001 |

<sup>a</sup>Adjusted for other variables in the table. <sup>b</sup>'Other' and 'Unknown' ethnic group categories not shown. <sup>c</sup>Unknown IMD quintiles not shown. <sup>d</sup>Unknown BMI not shown. IMD = Index of Multiple Deprivation, OR = odds ratio, BMI = body mass index, QOF = Quality and Outcomes Framework, ref = reference. Intraclass correlation coefficient for practice variation for both Model 1 and Model 2 is 0.16 (95% CI = 0.13 to 0.20)

A sensitivity analysis using individual co-morbidities, rather than numbers of LTCs did not improve the explanatory effect of the model. (see Table 1 supplementary data)

## Discussion

### Summary

Using patient level data from the GP record this study documents the numbers of suspected COVID-19 cases presenting to practices through the peak of the London epidemic (Figure 2). Data from these GP suspected cases illuminates predictors of infection at an earlier stage of the disease trajectory than data from hospital or ONS case fatality reports. (8, 14)

**Figure 2.**
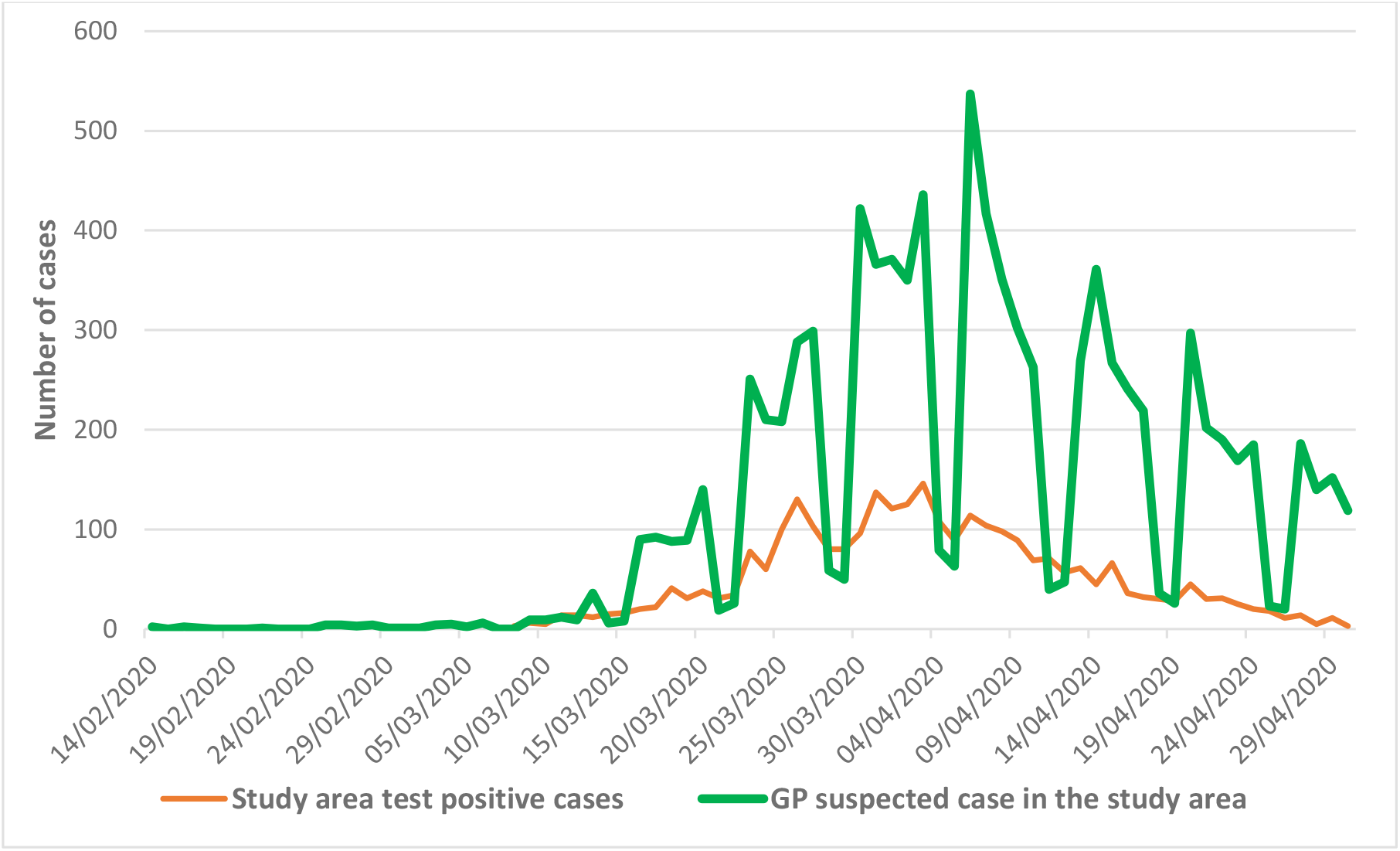
Comparison of study area test-positive cases and daily counts of GP suspected cases: 10 Feb 2020 to 30 April 2020.

The close to two-fold increase in the odds of infection for South Asian and Black groups shown in the univariate analysis (Table 1) is reduced by only a small amount when adjusted by demographic and clinical factors in the multivariate analysis (Table 2). The sizeable residual enhanced risk for ethnic minority groups in the fully adjusted analysis remains unexplained.

The number of co-morbidities in adult patients, and being overweight or obese are both major independent risk factors, but the overall effect of social deprivation was reduced in the multivariate analysis.

Figure 2 shows that GP coding for suspected COVID-19 follow the same distribution as test-positive cases, but with a threefold greater volume, reflecting the large number of community cases. Additional symptomatic individuals will have contacted NHS 111; with many others making no contact with health services – including those cases who were asymptomatic. Data on viral tests done either in community or hospital settings were not routinely available to general practice (12).

Figure 3 shows that recorded upper and lower respiratory infection episodes fell sharply during March, during the period with a rise in suspected COVID cases. This reflects the usual seasonal decline in viral URTIs, which may have been enhanced by social distancing. The national RCGP surveillance practice data show similar trends. (23, 24) This data suggests that GPs were distinguishing COVID symptoms from those of seasonal URTIs.

**Figure 3.**
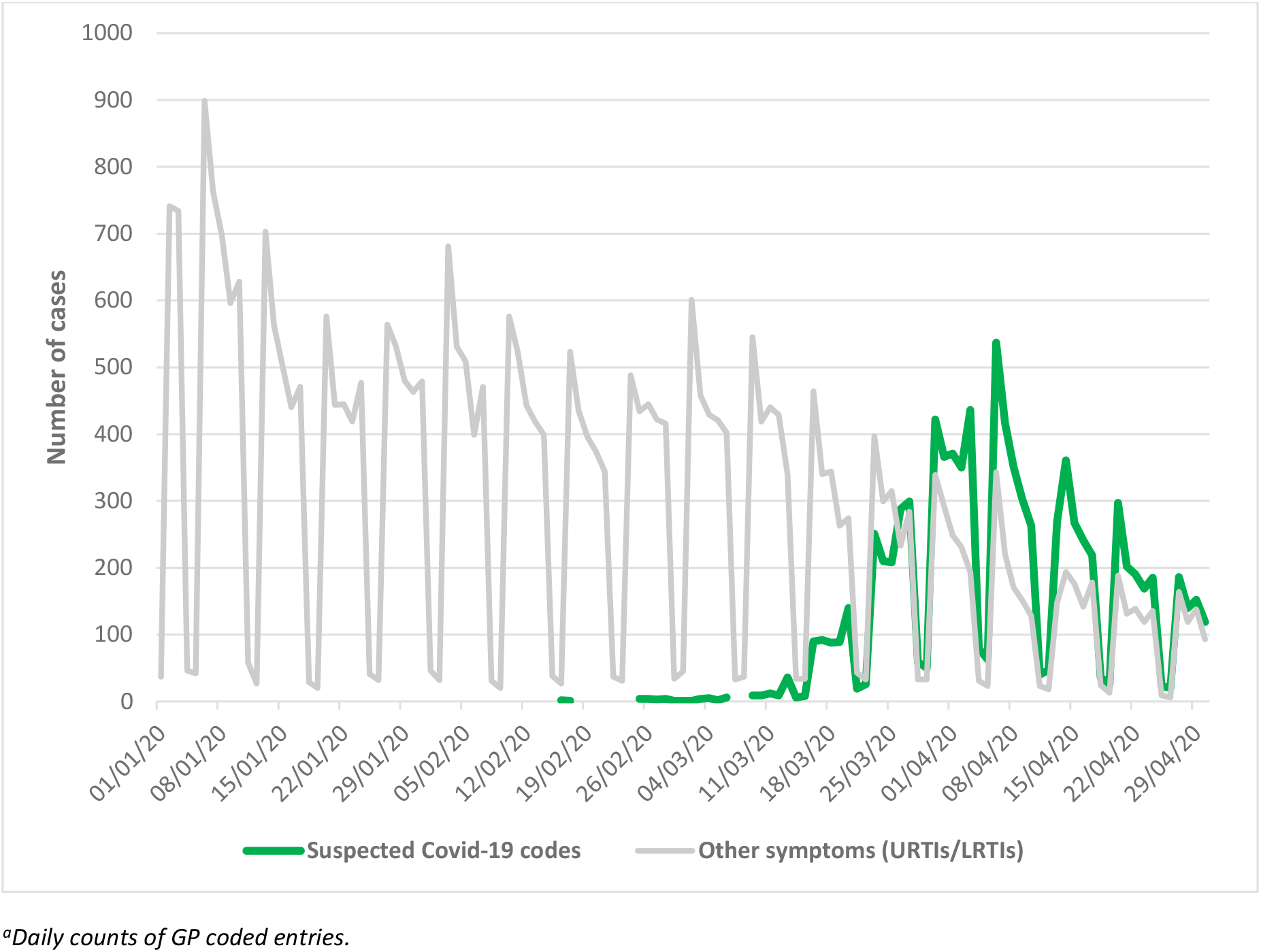
Comparison of GP suspected cases with GP URTI/LRTI codes across the study CCGs: 1 Jan 2020 to 30 April 2020^a^.

### Strengths and limitations

The strength of this study is based on the use of primary care data for the entire population registered at 157 general practices in adjacent CCGs in east London. The high level of ethnicity recording, coupled with the accurate recording of co-morbidities associated with the QOF, provides a unique opportunity to explore how clinical factors and demography affect the prevalence of suspected COVID-19 by ethnicity. Using UK government data on test-confirmed cases by London borough, (22) we confirm that GP coded data for suspected COVID-19 follows the same time course as the London epidemic (Figures 1,2). The inclusion of all episodes of URTI and LRTI from January suggest good separation of these clinical syndromes in east London practices. Data from RCGP surveillance practices suggest BAME populations present to GPs with URTI at similar rates to the white population. (24)

Limitations common to studies using routinely collected clinical data include potential diagnostic inaccuracies, and under-recording of some conditions. General practitioners did not have access to COVID-19 viral testing, hence the majority of recorded cases reflect suspected disease. It is likely that this report underestimates the effect size, there will be many asymptomatic, mildly ill, or patients who contacted NHS 111 (but not their practice) in the population not coded for suspected COVID-19. In contrast to studies which use an extended list of co-morbidities or weighted co-morbidity scores (25) we used a simple count of 16 conditions in the UK pay for performance QOF, as these are well recorded across practices.(20)

We were unable to include potentially important measures, such as household size and inter-generational composition, employment factors including travel and activity more likely to result in exposure, or the availability of personal protective equipment. Such social and cultural factors are likely to make significant contributions to the observed differences in disease prevalence by ethnicity, but may require bespoke data sets to provide answers.

### Comparison with existing literature

The trends in risk from this report are largely consistent with the findings on ethnicity, socioeconomic status and risk of death from COVID-19 based on hospital deaths, and with ONS reports which include deaths in hospital and community settings, adjusted by aggregate data on self-reported health and household composition, albeit collected in 2011. (26)

This similarity in risk of disease for BAME adults is surprising, in that this report includes milder episodes of disease, many among younger people and mostly managed within primary care. In contrast with other reports we did not see an excess of male cases. This may reflect a reluctance to consult, or that gender differences only become apparent further along the disease trajectory.

The risks of disease associated with smoking have been disputed, with some studies showing lower risks of positive tests, hospital admission or death among current smokers. (9, 27) A recent meta-analysis suggests higher risks for smokers and those with COPD. (28) The coded smoking data in our report was limited to current smoking/non-smoking-status. This may introduce bias, in that recent ex-smokers – who may stop because of respiratory symptoms or cardiovascular disease - are included among the non-smokers. Hence smoking was not included in the multivariate analysis.

### Implications for practice and research

This study demonstrates that much of the Covid-19 epidemic is being managed in primary care, which has rapidly adjusted to requirements for non face-to-face consultations.

Consultations in general practice may be useful as an early warning system for detection and monitoring of new outbreaks of disease which may follow the relaxation of lockdown restrictions. Practice infrastructure should be utilised to support testing and contact tracing. Ensuring the timely reporting of COVID-19 test results to practices, and diagnostic information from NHS 111, is a necessary part of this strategy, and will enable practices to provide continuing care to patients with more severe episodes.

Unpicking the underlying reasons for the higher risk of COVID-19 infection among those from ethnic minority populations will require studies which include data from a range of other sources, including household composition, overcrowding and a range of factors associated with occupational exposure.

## Funding

No project specific funding

### Ethical approval

Based on the NHS Health Research Authority Questionnaire (http://www.hra-decisiontools.org.uk/ethics/) Research Ethics approval was not required for this project as patient-level data are anonymised, and only aggregated patient data are reported in this study. All GPs in the participating east London practices consented to the use of their anonymised patient data for research and development for patient benefit.

### Data sharing

All data relevant to the study are included in the article.

### Author contribution

The study was designed by KB and SH. Data analysis was by CW. The report was written by SH with contributions from all authors.

## Data Availability

All data relevant to the study are included in the article

## Acknowledgements

The authors are grateful to the participating GPs for their cooperation, without which, such studies would be impossible. The authors wish to thank staff at CEG for supporting practices with guidance and data entry tools which support this project.

## Supplementary data

**Supplementary Table 1.** Multivariate model for predictors of COVID-19 for adults aged ≥ 18 years, replacing counts of LTCs with individual LTCs.

|  |  | OR<sup>a</sup> | 95% CI | P Value |
| --- | --- | --- | --- | --- |
| **Sex** | Male (ref) | 1.00 |  |  |
|  | female | 1.15 | (1.10 to 1.20) | <0.001 |
| **Age bands (years)** | 18-49 (ref) | 1.00 |  |  |
|  | 50-69 | 1.41 | (1.33 to 1.49) | <0.001 |
| | $\geq 69$ | 1.24 | (1.14 to 1.36) | <0.001 |
| **Ethnicity<sup>b</sup>** | White (ref) | 1.00 |  |  |
|  | South Asian | 2.00 | (1.86 to 2.10) | <0.001 |
|  | Black | 1.53 | (1.43 to 1.63) | <0.001 |
| **Internal IMD 2015 quintiles<sup>c</sup>** | 1 least deprived (ref) | 1.00 |  |  |
|  | 2 | 1.18 | (1.09 to 1.28) | <0.001 |
|  | 3 | 1.15 | (1.07 to 1.25) | <0.001 |
|  | 4 | 1.21 | (1.12 to 1.31) | <0.001 |
|  | 5 most deprived | 1.26 | (1.16 to 1.36) | <0.001 |
| **BMI<sup>d</sup>** | Normal weight (18.5 to <25) (ref) |  |  |  |
|  | Underweight (<18.5) | 0.82 | (0.71 to 0.95) | 0.009 |
|  | Overweight (25 to <30) | 1.35 | (1.30 to 1.43) | <0.001 |
|  | Obese (30 to <35) | 1.84 | (1.73 to 1.95) | <0.001 |
| | Morbidly Obese ($\geq 40$) | 2.40 | (2.17 to 2.66) | <0.001 |
| **Asthma** |  | 1.40 | (1.32 to 1.48) | <0.001 |
| **Atrial Fibrillation** |  | 1.38 | (1.20 to 1.58) | <0.001 |
| **Cancer** |  | 1.39 | (1.26 to 1.54) | <0.001 |
| **Coronary heart disease** |  | 1.13 | (1.02 to 1.25) | 0.017 |
| **Chronic kidney disease (3-5)** |  | 1.20 | (1.09 to 1.31) | <0.001 |
| **COPD** |  | 1.43 | (1.27 to 1.61) | <0.001 |
| **Dementia** |  | 3.69 | (3.20 to 4.27) | <0.001 |
| **Depression** |  | 1.52 | (1.44 to 1.61) | <0.001 |
| **Diabetes** |  | 1.43 | (1.34 to 1.52) | <0.001 |
| **Hypertension** |  | 1.16 | (1.09 to 1.23) | <0.001 |
| **Severe Mental Illness** |  | 1.02 | (0.90 to 1.17) | 0.732 |
| **Stroke TIA** |  | 1.19 | (1.05 to 1.36) | 0.007 |

<sup>a</sup>Adjusted for other variables in the table. <sup>b</sup>'Other' and 'Unknown' ethnic group categories not shown. <sup>c</sup>Unknown IMD quintiles not shown. <sup>d</sup>Unknown BMI not shown. IMD = Index of Multiple Deprivation, OR = odds ratio, BMI = body mass index, COPD = chronic obstructive pulmonary disease, TIA = transient ischaemic attack, QOF = Quality and Outcomes Framework, ref = reference. Intraclass correlation coefficient for practice variation is 0.16 (95% CI = 0.13 to 0.20)

